# The DiSCover Project: Protocol and Baseline Characteristics of a Decentralized Digital Study Assessing Chronic Pain Outcomes and Behavioral Data

**DOI:** 10.1101/2021.07.14.21260523

**Authors:** Jennifer L. Lee, Christian J. Cerrada, Mai Ka Ying Vang, Kelly Scherer, Caroline Tai, Jennifer L.A. Tran, Jessie L. Juusola, Christine N. Sang

## Abstract

**Background:** Chronic pain affects approximately 50 million adults in the United States and impacts mood, everyday functioning, and quality of life. The challenges of analgesic clinical trials and, therefore, the approval of new non-opioid analgesics, are based in part on a fundamental lack of understanding of those outcomes that are relevant to an individual’s overall functioning.

**Objectives:** To determine the behaviors and health outcomes associated with chronic pain. This manuscript presents an overview of the study design, baseline health and behavioral characteristics of our sample, and preliminary findings of how behavioral characteristics differ between individuals with and without chronic pain.

**Methods:** The study is a decentralized digital longitudinal cohort study of 10,036 individuals (5,832 with chronic pain [CP] and 4,204 with no chronic pain [NCP]), age 18 years or older, living in the United States. The study period was one year. Data were collected from wearable activity trackers and health or fitness mobile applications to capture passively collected behavioral data including steps, sleep, and heart rate. Patient-reported outcomes on mood and pain, including the BPI-SF, PHQ-9, and GAD-7, were collected at various timepoints during the study.

**Results:** The data suggest greater levels of depression and anxiety, lower quality of life, less physical activity, more variable sleep, and higher resting heart rate are associated with CP.

**Conclusions:** The longitudinal data from the larger study will yield substantial contributions to the body of literature in chronic pain, particularly in delineating relational and causal factors relevant to the impact of chronic pain, and potential development of a digital biomarker to assess and monitor patients’ everyday experience with chronic pain.

## Introduction

Chronic pain, typically defined as pain lasting longer than three months, presents a significant health burden affecting about 50 million adults in the United States and negatively impacts quality of life (Dahlhamer et al., 2018; Dueñas, Ojeda, Salazar, Mico, & Failde, 2016). Chronic pain can be associated with identifiable etiologies such as osteoarthritis and peripheral and central nervous system injuries, but many people suffer from chronic pain of an unknown etiology (Arnold et al., 2016). In addition, chronic pain is associated with psychiatric comorbidities such as anxiety and depression (Bair, Robinson, Katon, & Kroenke, 2003; Sareen, Cox, Clara, & Asmundson, 2005). Despite the high prevalence and impact of chronic pain, its pathology remains poorly understood and effective treatment options are limited (Green, Wheeler, LaPorte, Marchant, & Guerrero, 2002; Papageorgiou, Silman, & Macfarlane, 2002; Turk, 2002; Turk, Wilson, & Cahana, 2011), and some treatment options are problematic, as evidenced by the current opioid crisis (Gostin, Hodge, & Noe, 2017). Investigations have begun utilizing digital interventions for chronic pain (Bailey et al., 2020), and exercise and physical therapy have also been investigated and encouraged as treatments for chronic pain (Ambrose & Golightly, 2015; Casey et al., 2020). Despite these intervention development efforts, little is known about levels of activity and exercise engaged in by people with chronic pain. Given the increased prevalence of wearable activity trackers and the increasing availability of real world data (i.e., data collected outside of clinical trials in participants’ everyday lives), obtaining a nuanced understanding of the experience of chronic pain over time, across multiple data sources and diverse medical conditions, in real world settings is critical to inform the development and refinement of such interventions.

A number of studies have examined chronic pain outcomes longitudinally (Papageorgiou et al., 2002; Tay, Willcocks, & Chen, 2014; Walitt et al., 2011; Young, Amatya, Galea, & Khan, 2017); despite the widespread availability of wearable devices for tracking passively collected behavioral data, there have been limited studies investigating these types of data in chronic pain. One study using a digital sensor measured physical activity for participants with varying chronic pain conditions and found that pain intensity had an impact on overall physical activity over a 5- day period (Paraschiv-Ionescu, Perruchoud, Rutschmann, Buchser, & Aminian, 2016). Another study on advanced-stage cancer found that higher levels of pain were significantly correlated with decreased step count (Gresham et al., 2018), while a third study found no association of pain intensity with activity level for people with chronic low back pain (Huijnen et al., 2011). There are studies assessing physiological outcomes such as heart rate and sleep in chronic pain that indicate these variables play a role in the development and persistence of chronic pain (Finan, Goodin, & Smith, 2013; Naranjo-Hernández, Reina-Tosina, & Roa, 2020). However, many of these studies use clinical-grade sensors, such as actigraphs, and not commercially available devices intended for daily use by individuals collecting data in their everyday lives, limiting their generalizability. At the time of publication of this article, no studies have examined the associations between passively collected data from multiple data types and chronic pain outcomes using commercially available devices during the activities of an individual’s everyday life.

The 2011 Institute of Medicine report on chronic pain (Relieving Pain in America: A Blueprint for Transforming Prevention, Care, Education, and Research. Washington, DC, Institute of Medicine, 2011), states that “data are lacking on the prevalence, onset, course, impact, and outcomes of most common chronic pain conditions.” This report guided the development of the National Pain Strategy (Interagency Pain Research Coordinating Committee, National Institutes of Health/Department of Health and Human Services, 2016). In an effort to meet their call to fill this gap, we conducted a digital study that generates real world data from questionnaires, connected wearable devices and health apps, and other medical data to compile detailed information from individuals living with chronic pain. This protocol paper provides an overview of the study design, describes the decentralized digital operations of the protocol, and presents baseline health and behavioral characteristics of the sample.

## Methods

### Study Design

The DiSCover Project (**D**igital **S**ignals in **C**hronic **P**ain) is a one-year longitudinal cohort study aimed at evaluating daily activity behaviors in individuals with and without chronic pain and developing digital biomarkers for chronic pain severity, flare-ups, and quality of life. The study was conducted entirely online using the Achievement Studies platform (Evidation Health, Inc., San Mateo, CA). The study platform was used to screen and enroll participants, as well as collect and monitor data. The protocol was approved by the committee on research ethics at the Western Institutional Review Board (WIRB) and was conducted in accordance with the Declaration of the World Medical Association (www.wma.net). The study is registered with clinicaltrials.gov (NCT03421223).

### Recruitment and Screening

Achievement is a community of users in the U.S. ranging in age from 18 to 80+ years. Achievement community members can connect health or fitness apps (e.g., Fitbit, Garmin, MyFitnessPal) to the study platform and log daily activities (e.g., steps, sleep) to earn points that can be redeemed for monetary rewards. Achievement members are also invited to participate in various research opportunities and can access Achievement via the web and a mobile application (app).

Participants were sent offers to participate in Achievement and via social media advertising. Through the online study platform, potential participants completed a set of screener questions to assess their eligibility. Eligible participants were 18 years or older, read/spoke English, and resided in the United States. Participants were assigned to the Chronic Pain (CP) cohort based upon a self-reported diagnosis of chronic pain, defined as having chronic pain most days for at least three months, and rating their chronic pain as typically moderate to severe on the Numeric Pain Rating Scale (i.e., reported a pain score ≥ 4). Individuals who reported having chronic pain below 4 on the Numeric Pain Rating Scale were excluded from the study. Participants who met the inclusion criteria but denied having chronic pain were assigned to the No Chronic Pain (NCP) cohort. Recruitment was monitored for balanced cohort enrollment.

### Study Enrollment and Procedures

Once eligible participants electronically consented to participate via an online study platform, they completed a baseline survey, consisting of questions on medical history (including chronic pain), medications, quality of life, health care utilization, comorbidities, and treatment regimens. Participants were considered enrolled in the study after they completed the baseline assessment. Participants could then connect at least one eligible wearable activity tracker (i.e., Fitbit, Garmin, Under Armour, Apple Health, and Samsung Health) and at least one eligible health or fitness app (i.e., MyFitnessPal, Strava, MapMyRun, Moves, MapMyWalk, MapMyFitness, and Withings [formerly Nokia Health]) to their online Achievement profile.

### Study Procedures for Cohort 1

During the first two months of the study, all participants in the CP cohort were asked to complete short daily online surveys regarding any breakthrough pain they experienced in the past 24 hours. If they experienced breakthrough pain, they were asked follow-up questions regarding the duration, severity, and potential cause of pain, and information on medication usage. Participants in the CP cohort were also asked to complete monthly online surveys. See Table 1 for the data assessment schedule. Subgroups of the CP cohort also engaged in additional sub-studies and procedures; further details on the procedures of these sub-studies will be discussed in future manuscripts.

**Table 1.** Assessment schedule for monthly surveys.

| Variables | Month of Participation |  |  |  |
| --- | --- | --- | --- | --- |
|  | Baseline/0 | 1, 11 | 2, 4, 5, 7, 8, 10 | 3, 6, 9, 12 |
| Demographics | X |  |  |  |
| Medical history | X | X |  |  |
| Health care utilization | X |  |  | X |
| PHQ-9 (depression) | X |  |  | X |
| GAD-7 (anxiety) | X |  |  | X |
| BPI-SF (pain severity and interference) | X | X | X | X |
| Medication/non-medication therapies | X | X | X | X |
| Lifestyle changes | X | X | X | X |
| ACPA-QOLS (quality of life) | X | X | X | X |

### Study Procedures for Cohort 2

Participants in the NCP cohort were asked to complete monthly online surveys, assessing the same domains as individuals in the CP cohort (see Table 1), but did not complete daily surveys.

## Measures of Clinical Characteristics

### Brief Pain Inventory – Short Form (BPI-SF) (Cleeland, 2009)

The BPI-SF is a self-report questionnaire that asks participants about their pain and the impact of pain on their daily functioning (Mendoza, Mayne, Rublee, & Cleeland, 2006). Portions of the BPI-SF were asked each month and participants rated their pain on a scale of 0-”No pain” to 10-”Pain as bad as you can imagine” within the past week on four qualities: worst, least, average, and right now. Participants also rated the level of interference their pain has on a scale of 0-”Does not interfere” to 10-”Completely interferes” with regards to the following activities: general activity, mood, walking, work, relations with others, sleep, and enjoyment of life, an endpoint that has been recommended for use in all clinical studies of pain (Dworkin et al., 2005). For patients with pain caused by a variety of medical diagnoses, a pain rating greater than 4 on a 0 to 10 scale is considered to be unacceptable (Tubach et al., 2012).

### Patient Health Questionnaire - 9 (PHQ-9) (Kroenke, Spitzer, & Williams, 2001)

The PHQ-9 is a 9-item, self-report of depressive symptoms. It is a well-validated questionnaire that has been used in studies for people with chronic pain (Dobscha et al., 2009). Participants endorsed how much they had been bothered by symptoms over the past two weeks on a 4-point Likert scale from 0-”Not at all” to 3-”Nearly every day,” with a sum of scores ranging from 0 to 27, with higher scores indicating higher levels of depressive symptoms. Internal consistency was good, with Cronbach’s α = .87 for CP cohort and .88 for NCP cohort.

### Generalized Anxiety Disorder Screener - 7 (GAD-7) (Spitzer, Kroenke, Williams, & Löwe, 2006)

The GAD-7 is a 7-item self-report of generalized anxiety symptoms. It is a well-validated questionnaire that has been used in RCTs for people with chronic pain (Dear et al., 2018). Participants endorsed how much they had been bothered by symptoms on a 4-point Likert scale from 0-”Not at all sure” to 3-”Nearly every day,” with the sum of scores ranging from 0 to 21, with higher scores indicating higher anxiety symptoms. Internal consistency in these samples was excellent, with Cronbach’s α = .92 for CP cohort and .92 for NCP cohort.

### American Chronic Pain Association-Quality of Life Scale (ACPA-QOLS) (Association)

The ACPA-QOLS is a single-item self-report of functioning for people with pain that has been used in research for pain treatments (Saeed et al., 2014). Participants were instructed to select the statement that best described their QOL over the past month, on a scale from 0-”Non-functioning” to 10-”Normal quality of life,” which included anchored descriptions of abilities to engage in activities of daily living such as work and social activities. For example, a 6 is being able to “work/volunteer limited hours, take part in limited social activities on weekends.”

### Data from Wearable Activity Trackers

Participant-level behavioral variables outlined in Table 2 were calculated using daily step, sleep, and heart rate data from activity trackers. Retrospective data from the 90-day period prior to enrollment was used, and data was also prospectively collected for the duration of the study period (one year). For the purposes of this protocol paper, day-level variables such as daily steps and daily hours asleep were aggregated across days within the 90-day period prior to enrollment to create participant-level variables. Classification of low vs. high step days and low vs. high sleep days were predefined based on suggested cutoffs (Tudor-Locke, Johnson, & Katzmarzyk 2009; Watson et al., 2015). Daily resting heart rate and maximum heart rate were only available for Fitbit users, and were similarly aggregated to describe participants. Because participants could connect more than one activity tracker to the study, only data from the tracker type with the greatest number of valid days of step, sleep, or heart rate data was used for analysis.

**Table 2.**
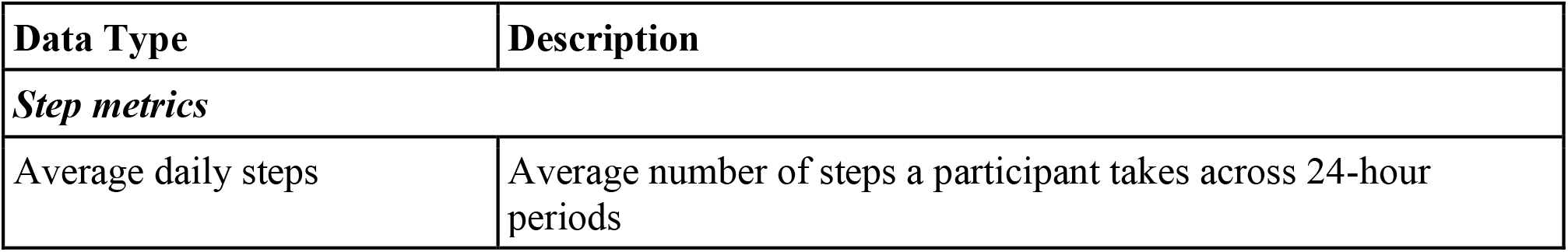
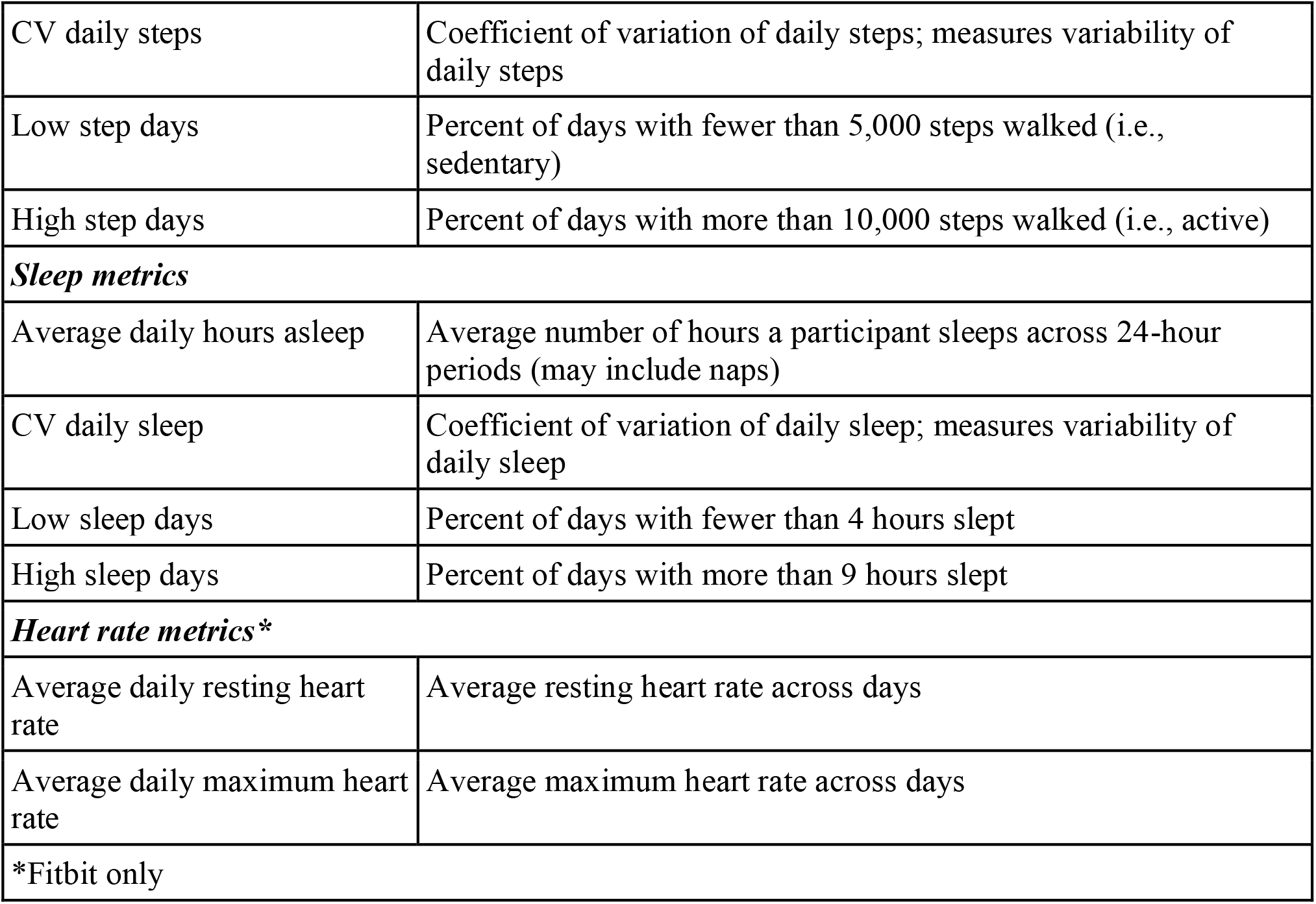
Participant-level behavioral variables.

### Statistical Analyses

Descriptive statistics (i.e., means, standard deviations, and percentages) were calculated for baseline demographic and clinical characteristics stratified by cohort (Table 3). We performed between-group comparisons (CP vs. NCP) to characterize cohorts with respect to clinical characteristics using appropriate statistical tests (e.g., Mann-Whitney U, t-test, or test of proportions). Between-group comparisons for activity data were also conducted for the per-patient behavioral variables outlined in Table 2. Extreme values were addressed by Winsorizing activity data at the 1^st^ and 99^th^ percentiles. Analysis was limited to the subset of participants who had activity data for at least 30 of the 90 days prior to study enrollment. This threshold was set to ensure that any behavioral characteristics or patterns inferred from tracker-based behavioral data were based on a robust set of participant data rather than sparse data points. For all statistical comparisons, p-values were corrected for False Discovery Rate (FDR) in order to minimize the Type 1 error rate due to multiple comparisons. For exploratory purposes, descriptive statistics for clinical characteristics and behavioral variables were additionally calculated for individuals within the CP cohort, stratified by the most commonly reported chronic pain conditions. Statistical analyses were conducted in R version 3.6.1 (R Foundation for Statistical Computing, Vienna, Austria).

**Table 3.**
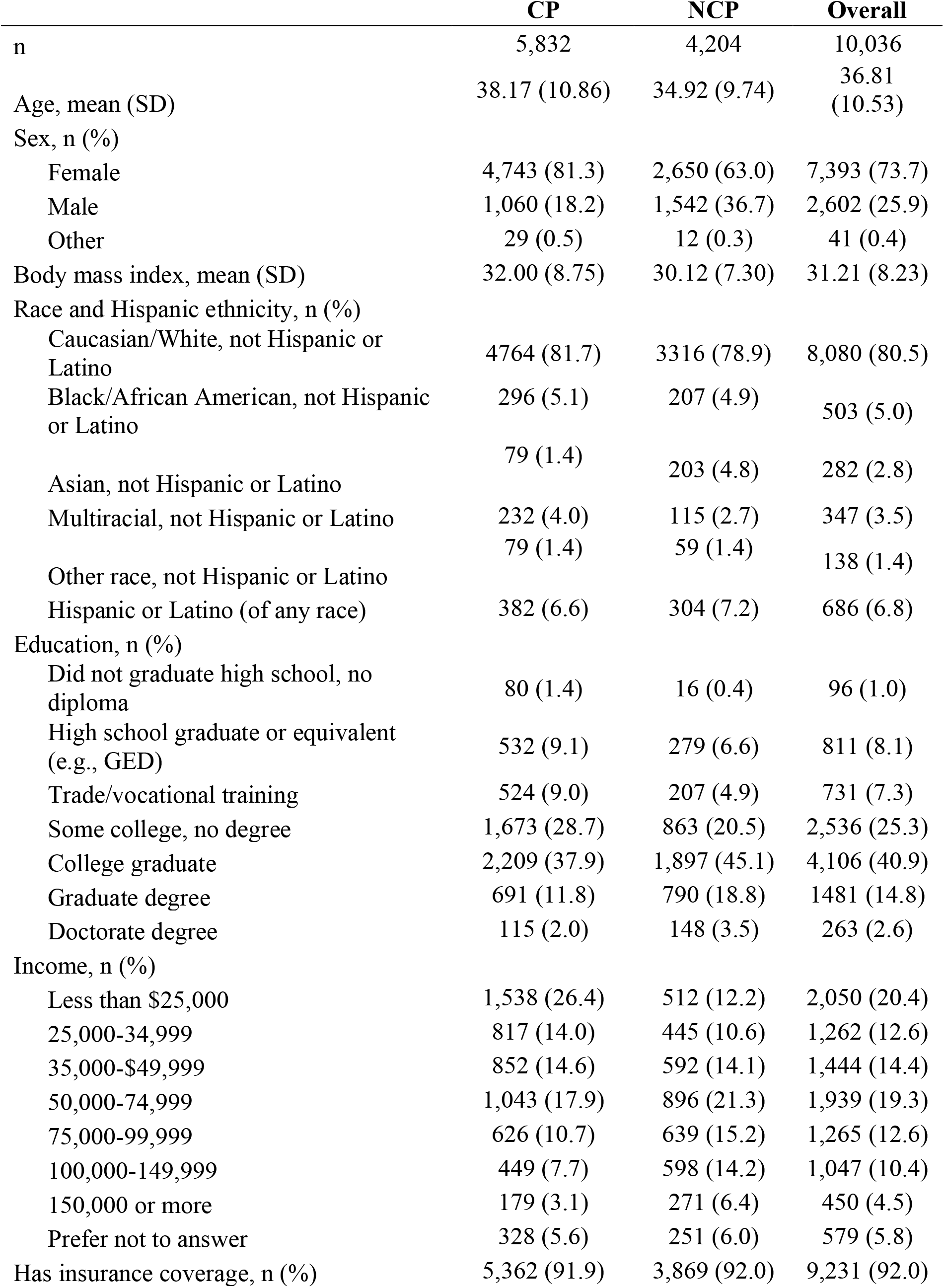
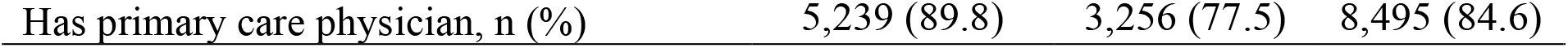
Demographic characteristics.

|  | **CP** | **NCP** | **Overall** |
| --- | --- | --- | --- |
| n | 5,832 | 4,204 | 10,036 |
| Age, mean (SD) | 38.17 (10.86) | 34.92 (9.74) | 36.81 (10.53) |
| Sex, n (%) |  |  |  |
| Female | 4,743 (81.3) | 2,650 (63.0) | 7,393 (73.7) |
| Male | 1,060 (18.2) | 1,542 (36.7) | 2,602 (25.9) |
| Other | 29 (0.5) | 12 (0.3) | 41 (0.4) |
| Body mass index, mean (SD) | 32.00 (8.75) | 30.12 (7.30) | 31.21 (8.23) |
| Race and Hispanic ethnicity, n (%) |  |  |  |
| Caucasian/White, not Hispanic or Latino | 4764 (81.7) | 3316 (78.9) | 8,080 (80.5) |
| Black/African American, not Hispanic or Latino | 296 (5.1) | 207 (4.9) | 503 (5.0) |
| Asian, not Hispanic or Latino | 79 (1.4) | 203 (4.8) | 282 (2.8) |
| Multiracial, not Hispanic or Latino | 232 (4.0) | 115 (2.7) | 347 (3.5) |
| Other race, not Hispanic or Latino | 79 (1.4) | 59 (1.4) | 138 (1.4) |
| Hispanic or Latino (of any race) | 382 (6.6) | 304 (7.2) | 686 (6.8) |
| Education, n (%) |  |  |  |
| Did not graduate high school, no diploma | 80 (1.4) | 16 (0.4) | 96 (1.0) |
| High school graduate or equivalent (e.g., GED) | 532 (9.1) | 279 (6.6) | 811 (8.1) |
| Trade/vocational training | 524 (9.0) | 207 (4.9) | 731 (7.3) |
| Some college, no degree | 1,673 (28.7) | 863 (20.5) | 2,536 (25.3) |
| College graduate | 2,209 (37.9) | 1,897 (45.1) | 4,106 (40.9) |
| Graduate degree | 691 (11.8) | 790 (18.8) | 1481 (14.8) |
| Doctorate degree | 115 (2.0) | 148 (3.5) | 263 (2.6) |
| Income, n (%) |  |  |  |
| Less than \$25,000 | 1,538 (26.4) | 512 (12.2) | 2,050 (20.4) |
| 25,000-34,999 | 817 (14.0) | 445 (10.6) | 1,262 (12.6) |
| 35,000-\$49,999 | 852 (14.6) | 592 (14.1) | 1,444 (14.4) |
| 50,000-74,999 | 1,043 (17.9) | 896 (21.3) | 1,939 (19.3) |
| 75,000-99,999 | 626 (10.7) | 639 (15.2) | 1,265 (12.6) |
| 100,000-149,999 | 449 (7.7) | 598 (14.2) | 1,047 (10.4) |
| 150,000 or more | 179 (3.1) | 271 (6.4) | 450 (4.5) |
| Prefer not to answer | 328 (5.6) | 251 (6.0) | 579 (5.8) |
| Has insurance coverage, n (%) | 5,362 (91.9) | 3,869 (92.0) | 9,231 (92.0) |
| Has primary care physician, n (%) | 5,239 (89.8) | 3,256 (77.5) | 8,495 (84.6) |

## Results

### Study Participants

Study recruitment, screening, and enrollment occurred between March and December of 2018. Approximately 25,000 participants completed a screener to determine eligibility (Figure 1- CONSORT diagram). A total of 10,036 individuals (5,832 in CP cohort and 4,204 in NCP cohort) completed informed consent and the online baseline assessment, and were enrolled in the study, a sample size that is comparable to other published studies on chronic pain (Bailey et al., 2020; Blyth et al., 2001; Yongjun et al., 2020). Of the individuals enrolled in the study, 8,899 (88.7%) connected at least one activity tracker to the Achievement Studies platform and 5.4% had connected at least one health or fitness app to the platform. Figure 2 depicts the geographic distribution of participants in the U.S., based upon ZIP code data, with representation from all 50 states, D.C., Puerto Rico, and the U.S. Armed Forces.

**Figure 1.**
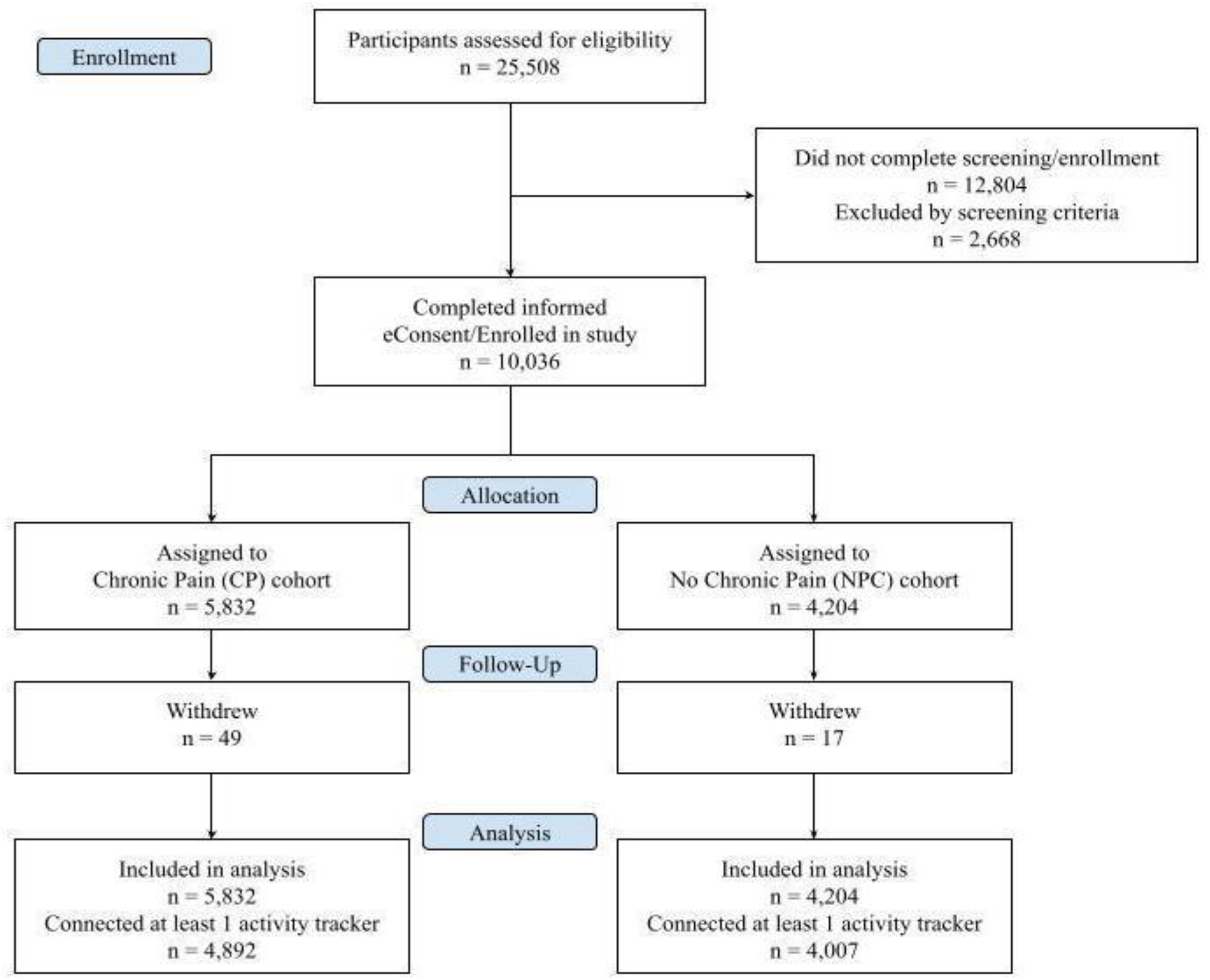
CONSORT diagram.

**Figure 2:**
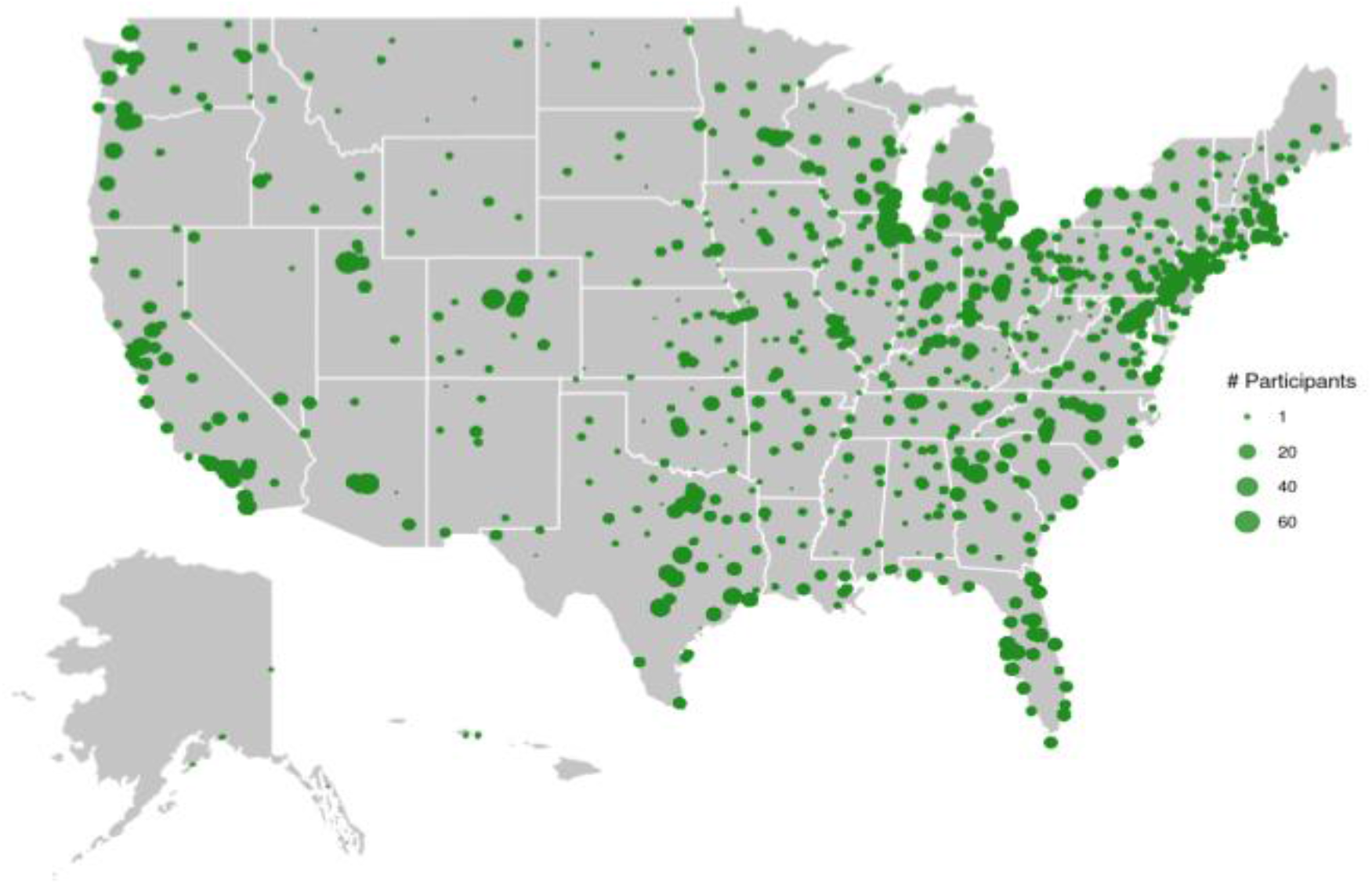
Geographic distribution map of study participants.

### Demographic and Clinical Characteristics

Table 3 presents demographic characteristics overall and by cohort. Overall, the majority of participants were young (18-35) and middle-aged adults (36-55; Mean=37.3; SD=10.5, range: 18 to 85) and primarily Non-Hispanic White (80.5%). A greater percentage of females were in the CP cohort compared to the NCP cohort (81.3% vs. 63.0%). Among CP participants, the most common self-reported conditions were migraine (61.8%), osteoarthritis (43.1%), fibromyalgia (34.0%), and peripheral nerve pain (26.0%) (See Appendix for complete list of conditions). The majority of NCP participants did not report any of the previous conditions from the list presented (68.2%), and 19.4% of them reported experiencing migraine. Individuals with CP had been diagnosed with their condition for an average of 9.2 years (SD=7.2). Baseline medication use among CP participants was common; 4,554 (78.1%) were currently using OTC pain relief or patches and 3,667 (62.9%) were taking prescription medication, over a third of which (1,497, 40.8%) were prescription opioids. Approximately 44% of the CP cohort reported the use of digital apps for meditation, diet, or exercise.

Between-group comparisons of clinical characteristics (Table 4) showed that mean pain ratings were higher in the CP cohort compared to the NCP cohort by 2.9 points on severity and 3.5 points on interference (p’s<.001). The CP cohort also had higher mean PHQ-9 and GAD-7 scores and lower quality of life (p’s<0.001).

**Table 4.** Clinical characteristics.

|  | CP | NCP | Overall |
| --- | --- | --- | --- |
|  | 5,832 | 4,204 | 10,036 |
| BPI-Severity, mean (SD) | 5.25 (1.49)* | 2.38 (1.77) | 4.05 (2.15) |
| BPI-Interference, mean (SD) | 5.62 (2.09)* | 2.08 (2.11) | 4.14 (2.73) |
| PHQ-9 Score, mean (SD) | 11.91 (6.17)* | 6.34 (5.46) | 9.58 (6.49) |
| GAD-7 Score, mean (SD) | 8.98 (5.95)* | 5.15 (5.04) | 7.38 (5.90) |
| ACPA-QOLS in past month, mean (SD) | 6.33 (2.70)* | 8.54 (1.96) | 7.26 (2.65) |

\* $p < .001$ , FDR corrected p-value for multiple comparisons

### Activity Tracker-Based Behavioral Characteristics

Of the 8,899 participants with at least one connected activity tracker or app, 8,100 (91%) had dense daily step data (at least 30 days in the 90-day pre-enrollment period), the majority of which were collected from Fitbit (4,821, 60%) and Apple Health (2,106, 26%). Additionally, 4,859 (55%) participants had dense daily sleep data, the majority of which were from Fitbit (4,184, 86%). The remainder of step and sleep data came from trackers and apps such as Garmin, Samsung Health, Under Armour, and Withings. Finally, 3,725 (42%) participants had dense resting and maximum heart rate data collected from Fitbit. Among individuals with dense activity data, coverage was high; the average number of days with valid data in the 90-day pre-enrollment period across all participants were 74.7, 69.7, and 77.2 for step, sleep, and heart rate, respectively.

Overall, individuals in the CP cohort were less active and had less consistent sleep patterns. On average, CP individuals walked 2,209 fewer steps per day and had greater daily step variability, more sedentary days (5.4% vs. 2.2%), and less active days (19.6% vs. 33.9%) compared to NCP individuals (all p’s<.001). Compared to the NCP cohort, CP participants additionally had less regular sleep as measured by coefficient of variation. Furthermore, CP participants had greater percentages of both low and high sleep days (p’s<.001), which additionally suggest irregular sleep patterns. Similar differences in activity were observed for the subgroup of participants who were Fitbit users, except for average daily hours asleep (p=.74). Finally, among Fitbit users, CP participants had greater average daily resting heart rate than NCP participants by over 4 beats per minute (p<.001).

### Clinical Characteristics and Activity Tracker-Based Behavioral Characteristics within the Chronic Pain Cohort

Table 6 presents descriptive summaries of the clinical assessments stratified by chronic pain condition at baseline. Migraine, fibromyalgia, neuropathic pain, and arthritis/osteoporosis were the most common conditions in the CP cohort. All other conditions were combined. Within the CP cohort, participants with neuropathic pain reported the greatest pain and interference, and lowest quality of life relative to people with chronic pain who had none of the selected conditions.

**Table 5.** Behavioral characteristics.

|  | All activity trackers |  | Fitbit trackers only |  |
| --- | --- | --- | --- | --- |
|  | CP | NCP | CP | NCP |
| **Walk metrics** |  |  |  |  |
| n | 4,314 | 3,786 | 2,168 | 2,653 |
| Average daily steps, mean (SD) | 6,354.24<br>(3,853.04)* | 8,563.16<br>(3,889.68) | 7,409.03<br>(3,608.49)* | 9,141.24<br>(3,527.50) |
| CV daily steps | 0.56* | 0.48 | 0.46* | 0.43 |
| Low step days | 5.4%* | 2.2% | 2.1%* | 1.2% |
| High steps days | 19.6%* | 33.9% | 24.8%* | 37.8% |
| **Sleep metrics** |  |  |  |  |
| n | 2,357 | 2,502 | 1,915 | 2,269 |
| Average daily hours asleep, mean (SD) | 6.76 (1.16)* | 6.65 (0.95) | 6.53 (1.05) | 6.55 (0.89) |
| CV daily hours asleep | 0.26* | 0.23 | 0.26* | 0.23 |
| Low sleep days | 9.8%* | 7.6% | 10.9%* | 8.0% |
| High sleep days | 11.3%* | 7.1% | 8.4%* | 5.8% |
| **Heart rate metrics** |  |  |  |  |
| n | - | - | 1,680 | 2,045 |
| Average daily resting heart rate, mean bpm (SD) | - | - | 70.18 (8.51)* | 66.03 (7.90) |
| Average daily maximum heart rate, mean bpm (SD) | - | - | 104.35 (8.08)* | 103.55 (7.92) |

\* p<.001, FDR corrected p-value for multiple comparisons

**Table 6.** Clinical characteristics within Chronic Pain Cohort.

|  | **Chronic Pain Subgroups by Conditions in the Case Arm** |  |  |  |  |
| --- | --- | --- | --- | --- | --- |
|  | Migraine<br>only<br>(n=849) | Fibromyalgia<br>only<br>(n=220) | Neuropathic pain<br>only<br>(n=190) | Arthritis/osteoporosis<br>only<br>(n=518) | None of the<br>conditions<br>(n=714) |
| BPI-Severity, mean<br>(SD) | 4.9 (1.5) | 4.9 (1.2) | 5.4 (1.6) | 5.1 (1.6) | 4.7 (1.4) |
| BPI-Interference,<br>mean (SD) | 5.1 (2.0) | 5.3 (1.9) | 5.7 (2.1) | 5.2 (2.2) | 4.6 (2.0) |
| PHQ-9 Score, mean<br>(SD) | 11.6 (6.1) | 11.9 (5.9) | 11.6 (6.3) | 9.6 (6.0) | 10.0 (6.0) |
| GAD-7 Score, mean<br>(SD) | 9.5 (5.8) | 8.9 (5.8) | 8.8 (5.8) | 7.1 (5.6) | 7.8 (5.6) |
| ACPA-QOLS in past<br>month, mean (SD) | 6.7 (2.7) | 6.6 (2.7) | 6.1 (2.8) | 6.8 (2.7) | 7.4 (2.4) |

Behavioral characteristics were also described within the CP cohort stratified by conditions (Table 7). Within the CP cohort, individuals with fibromyalgia only walked the least steps per day, followed by people with migraine only. Average daily hours asleep were lowest in people with neuropathic pain, but sleep was fairly similar across groups. Average resting daily heart rate was highest in individuals with fibromyalgia only, followed by individuals with migraine only.

**Table 7.**
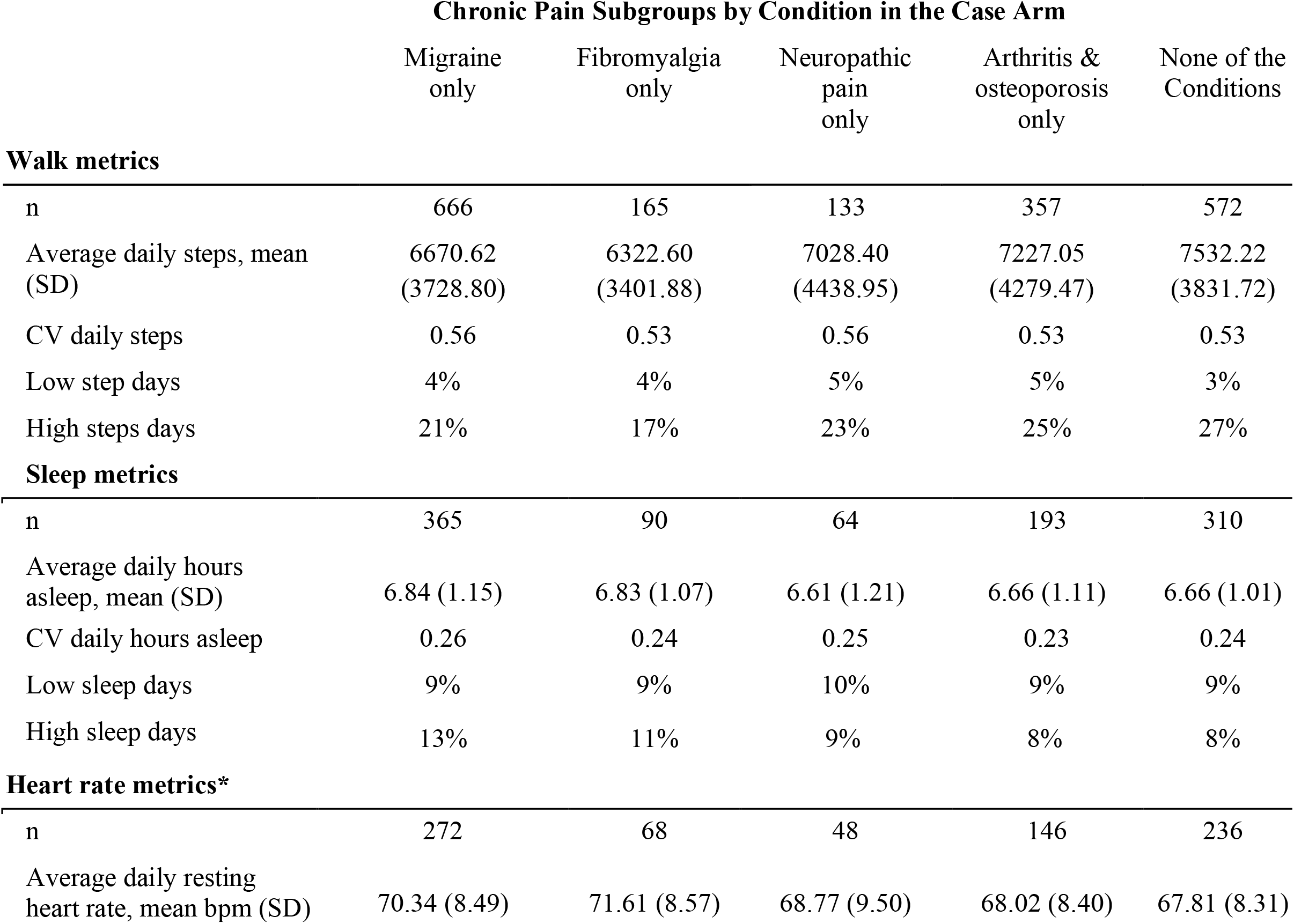
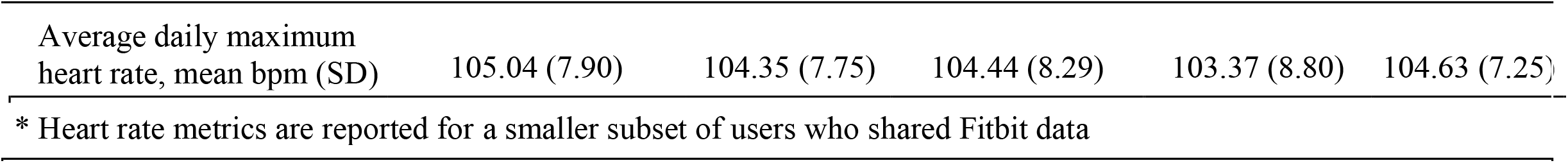
Behavioral characteristics within Chronic Pain Cohort.

|  | **Chronic Pain Subgroups by Condition in the Case Arm** |  |  |  |  |
| --- | --- | --- | --- | --- | --- |
|  | Migraine only | Fibromyalgia only | Neuropathic pain only | Arthritis & osteoporosis only | None of the Conditions |
| **Walk metrics** |  |  |  |  |  |
| n | 666 | 165 | 133 | 357 | 572 |
| Average daily steps, mean (SD) | 6670.62 (3728.80) | 6322.60 (3401.88) | 7028.40 (4438.95) | 7227.05 (4279.47) | 7532.22 (3831.72) |
| CV daily steps | 0.56 | 0.53 | 0.56 | 0.53 | 0.53 |
| Low step days | 4% | 4% | 5% | 5% | 3% |
| High steps days | 21% | 17% | 23% | 25% | 27% |
| **Sleep metrics** |  |  |  |  |  |
| n | 365 | 90 | 64 | 193 | 310 |
| Average daily hours asleep, mean (SD) | 6.84 (1.15) | 6.83 (1.07) | 6.61 (1.21) | 6.66 (1.11) | 6.66 (1.01) |
| CV daily hours asleep | 0.26 | 0.24 | 0.25 | 0.23 | 0.24 |
| Low sleep days | 9% | 9% | 10% | 9% | 9% |
| High sleep days | 13% | 11% | 9% | 8% | 8% |
| **Heart rate metrics\*** |  |  |  |  |  |
| n | 272 | 68 | 48 | 146 | 236 |
| Average daily resting heart rate, mean bpm (SD) | 70.34 (8.49) | 71.61 (8.57) | 68.77 (9.50) | 68.02 (8.40) | 67.81 (8.31) |
| Average daily maximum<br>heart rate, mean bpm (SD) | 105.04 (7.90) | 104.35 (7.75) | 104.44 (8.29) | 103.37 (8.80) | 104.63 (7.25) |

\* Heart rate metrics are reported for a smaller subset of users who shared Fitbit data

## Discussion

The novelty of the study design lies in its pairing of self-reported outcomes and passively collected behavioral data to better understand the daily lives of individuals with and without chronic pain over time in real-world settings. Analysis of the baseline data confirmed that participants with chronic pain experienced greater symptoms of depression and anxiety than individuals without chronic pain. These results are consistent with other research on chronic pain, suggesting this unique, digitally-recruited sample is similar to others with chronic pain (Bair et al., 2003; Sareen et al., 2005). Participants with chronic pain also reported lower quality of life compared to participants without chronic pain, consistent with current literature that chronic pain negatively impacts individuals’ quality of life (Dueñas et al., 2016).

Demographic analysis of the sample reveals differences in chronic pain by income level; a greater proportion of participants with lower incomes reported experiencing chronic pain when compared to participants with higher incomes. Chronic pain can be disabling and limit occupational opportunities, and people with lower incomes in the U.S. disproportionately experience chronic pain (Johannes, Le, Zhou, Johnston, & Dworkin, 2010). Participants with lower income may also have less access to and fewer opportunities for treatment of both acute and chronic pain, potentially resulting in greater levels of chronic pain. Future longitudinal analysis from this study may be beneficial in elucidating how demographic factors influence the experience of pain within this sample.

A greater number of women than men in the U.S. generally experience chronic pain, consistent with the greater numbers of females in the CP cohort in this study (Dahlhamer et al., 2018). Notably, the distribution by sex in the NCP cohort was slightly more female compared to the U.S. population, but was significantly lower than the CP cohort. The majority of participants were also young to middle-aged and non-Hispanic White, which raises the possibility that the findings may not be generalizable to other populations of individuals who experience chronic pain. A higher prevalence of chronic pain is usually associated with advancing age (Dahlhamer et al., 2018), and African Americans, in particular, have reported greater perceived pain severity and more interference with daily functions due to their chronic pain (Campbell & Edwards, 2012; Ruehlman, Karoly, & Newton, 2005). Despite the primarily non-Hispanic White sample, the number of individuals from diverse backgrounds in each group are sufficiently large to allow for future analyses with these specific subgroups. Future research should target a more diverse sample to further understand the experiences of these specific groups.

Analysis of behavioral data from this sample indicates significant differences in health-related behaviors between the CP and NCP cohorts at baseline. Overall, participants with chronic pain were less active on all metrics of physical activity. Participants with chronic pain also experienced poorer sleep, including less consistency in sleep patterns. However, differences in total sleep hours were not found between the CP and NCP cohorts when limiting to Fitbit users only, suggesting that some of the differences may also be driven by variability or factors related to wearable type. It is well-established that sleep and chronic pain have a complex relationship, the nuances and interrelatedness of which are still to be established in different populations (Finan et al., 2013). Among those with heart rate data available, individuals with chronic pain had a higher average daily resting heart rate. Research has suggested heart rate may play a role in the presentation and persistence of chronic pain, but literature is limited to smaller samples, only populations that present for medical care, or short follow-up due to the burden of collecting this type of data via electrocardiogram (Naranjo-Hernández et al., 2020). Within the CP cohort, initial descriptive statistics seem to suggest that there may be differences between individuals who experience chronic pain associated with migraine and fibromyalgia, and this requires further investigation in the longitudinal dataset. For example, further analysis of this data will be beneficial in connecting activity, sleep, and heart rate changes to instances of breakthrough pain and whether these associations are stronger for certain medical conditions.

This study has strengths, as well as limitations. The study enrolled over 10,000 participants who were geographically distributed across the U.S., suggesting it is feasible to engage large populations in decentralized digital clinical trials to better understand the behaviors and health outcomes of individuals with and without chronic pain. Furthermore, the study enrolled large subgroups of individuals with specific diseases like osteoarthritis and fibromyalgia, aligning with current literature that individuals with pain usually have one or more comorbid pain and/or chronic health conditions (Davis, Robinson, Le, & Xie, 2011). Despite their diversity of conditions, participants were recruited online, so they represent a digitally connected population that is interested in health behavior. Generalization to less connected populations may be limited. This study also had a healthy comparison control group to account for confounding variables that could explain the observed differences. Participants who wore Fitbits represented the largest proportion of participants within both groups, and contributed unique types of data (i.e., sleep and heart rate). It is possible that there were differences in measurement between wearable device type that are currently unaccounted for in this analysis, or characteristics of individuals who purchase a Fitbit over another wearable device that may be associated with their levels of physical activity and sleep. Further research is needed in this novel research area.

## Conclusion

The data from this initial baseline assessment and retrospective analysis suggest that this study has captured a relevant cohort of participants with and without chronic pain. The longitudinal behavioral, wearable, and clinical data has the potential to yield substantial contributions to the body of literature in chronic pain, particularly in delineating relational and causal factors relevant to the impact of chronic pain in everyday life. Given the year-long follow-up period and large sample size, a number of critical research questions surrounding pain can be addressed, including the disentanglement of the temporal relationships between pain, mood, and quality of life.

This cohort study includes multiple medical diagnoses in people with pain, which will also allow for the examination of differences between the experience of chronic pain across conditions. Condition-specific longitudinal analyses can take into consideration the natural disease course of conditions that typically are more chronic, chronic conditions that experience flares or episodic symptom exacerbations, or those that are typically more episodic. Longitudinal data on medication usage and types will also be beneficial for the examination of patterns of medication usage in relationship to pain symptoms, mood, and quality of life. The inclusion of activity data in these analyses would also help to assess whether and how medication usage or type may impact everyday functioning and metrics, such as steps, sleep, and heart rate.

Data from this longitudinal cohort study will help to quantify changes in activity data, such as steps, sleep, and heart rate, that occur when someone experiences a change in their symptoms and functioning, serving as an informative passive tool for health care providers. The development of a digital profile and biomarkers for chronic pain using passively generated data has been called for in the literature, but is not yet available and validated (Naranjo-Hernández et al., 2020). Digital profiles created from these biomarkers have potential to identify cohorts of individuals that may benefit from specific types of interventions or medications. Biomarkers from wearable data could also be used to assess and monitor response to treatment proactively in clinical practice, and better understand the patient’s everyday experience with chronic pain.

## Data Availability

Participant-level data will not be made available to the public; only aggregate results will be shared. The following documents will be provided and made available depending on availability to qualified researchers: Protocol, survey questionnaire, Statistical Methods, Final results presentation. Requests should be directed to to gain access to other data.

## Acknowledgments

The authors would like to thank Shefali Kumar and Claire Meunier for leading the conceptualization and design of the study; Katie Ulhorn for participant support; Lauren Vucovich for product and engineering support; Jermyn See for data preparation support; and members of the Achievement Studies Platform for their participation in this research study and time commitment to engaging in research activities.

## Appendix

**Table 9.** Comorbid conditions.

|  | **CP** | **NCP** | **Overall** |
| --- | --- | --- | --- |
| n | 5,832 | 4,204 | 10,036 |
| Comorbid conditions, n (%) |  |  |  |
| Cancer | 366 (6.3) | 84 (2.0) | 450 (4.5) |
| Diabetes, Type 1 | 79 (1.4) | 36 (0.9) | 115 (1.1) |
| Diabetes, Type 2 | 605 (10.4) | 170 (4.0) | 775 (7.7) |
| Fibromyalgia | 1981 (34.0) | 102 (2.4) | 2083 (20.8) |
| Gout | 240 (4.1) | 75 (1.8) | 315 (3.1) |
| Migraine | 3603 (61.8) | 814 (19.4) | 4417 (44.0) |
| Multiple sclerosis | 125 (2.1) | 21 (0.5) | 146 (1.5) |
| Peripheral neuropathic pain | 1519 (26.0) | 94 (2.2) | 1613 (16.1) |
| Central neuropathic pain | 445 (7.6) | 19 (0.5) | 464 (4.6) |
| Osteoarthritis | 2511 (43.1) | 441 (10.5) | 2952 (29.4) |
| Osteoporosis | 267 (4.6) | 31 (0.7) | 298 (3.0) |
| Rheumatoid arthritis | 698 (12.0 ) | 68 (1.6) | 766 (7.6) |

